# Havana Syndrome Among Canadian Diplomats: Brain Imaging Reveals Acquired Neurotoxicity

**DOI:** 10.1101/19007096

**Authors:** Alon Friedman, Cynthia Calkin, Amanda Adams, Guillermo Aristi Suarez, Tim Bardouille, Noa Hacohen, A. Laine Green, R. Rishi Gupta, Javeria Hashmi, Lyna Kamintsky, Jong Sung Kim, Robert Laroche, Diane MacKenzie, Dan Milikovsky, Darren Oystreck, Jillian Newton, Greg Noel, Jonathan Ofer, Maher Quraan, Claire Reardon, Margaux Ross, Derek Rutherford, Matthias Schmidt, Yonatan Serlin, Crystal Sweeney, Janine Verge, Leah Walsh, Chris Bowen

## Abstract

**BACKGROUND:** In late 2016, US diplomats stationed in Havana began presenting with a variety of neurological manifestations that proved difficult to diagnose. Though previous studies suggested a likely association with brain injury, the mechanism of injury, brain regions involved, and etiology remained unknown.

**METHODS:** We conducted a multimodal study examining 26 Canadian diplomats and their family members, the majority of whom presented with symptoms similar to their American counterparts while residing in Havana. Assessments included a medical history, self-reported symptom questionnaires, cognitive assessments, blood tests, and brain imaging assessments (magnetic resonance imaging (MRI) and magnetoencephalography (MEG)). Individuals showing signs of brain injury underwent further neurological, visual, and audio-vestibular assessments. Eight participants were tested both before and after living in Havana.

**RESULTS:** Our assessment documents multiple functional and structural impairments, including significant spatial memory impairment, abnormal brain-stem evoked potentials, degradation of fibre tracts in the fornix and posterior corpus callosum, blood-brain barrier injury to the right basal forebrain and anterior insula, and abnormal paroxysmal slowing events of cortical activity. Subsequent mass-spectrometry and blood analyses documented reduced serum cholinesterase activity and the presence of organophosphates (Temephos) and pyrethroid metabolites (3-phenoxybenzoic acid or 3-BPA).

**CONCLUSIONS:** Our findings confirm brain injury, specify the regions involved, and raise the hypothesis of overexposure to cholinesterase inhibitors as a plausible etiology. If correct, our hypothesis bears public health ramifications (see Discussion) and suggests a course of action for reducing exposure in the future.

**FUNDING:** Global Affairs Canada.

## Introduction

In autumn 2016, US diplomats in Havana began reporting unusual sensory and auditory stimuli accompanied by symptoms of dizziness, tinnitus, and various cognitive manifestations. Initial testing revealed abnormalities in cervical and ocular vestibular evoked myogenic potential metrics and were interpreted as a peripheral vestibular pathology affecting the otolithic organs.^1^

When 24 of the original subjects were retested approximately 200 days after the suspected exposure, clinical findings included cognitive, vestibular, and oculomotor abnormalities and suggested “a sustained injury to widespread brain networks.”^2^

To further understand the nature of injury and brain regions involved, we conducted multimodal clinical and quantitative imaging assessments for brain injury among 26 Canadian diplomats and their families stationed in Havana, the majority of whom presented with symptoms similar to their American counterparts in the months following autumn 2016.

## Methods

This was a multimodal, longitudinal, and quantitative study to examine behavioural and physiological indices of acquired brain injury. The study was approved by the Research Ethics Board of the Nova Scotia Health Authority. All participants provided written informed consent. Clinical tests were performed according to guidelines (see Appendix for more information).

### EXPOSED & NON-EXPOSED SUBJECTS

Between August 10, 2018, and February 20, 2019, we tested 26 Canadian adult subjects referred by Global Affairs Canada for evaluation. Of this group, 23 were considered “exposed” while 12 were considered “non-exposed,” with double testing (pre- and post-exposure) accounting for overlap between the two groups (see below).

### RECENTLY EXPOSED & REMOTELY EXPOSED

To better understand the progression of injury over time, data sets were separated according to when we were able to test our subjects. Within the exposed group, we were able to test 11 individuals within one month of their return from Havana, a group we classified as “recently exposed,” and 12 individuals 1-19 months after returning (median 14 months), a group we classified as “remotely exposed,” with double testing between these two subgroups accounting for overlap in figures (see below).

### REPEATED TESTING

Some subjects were tested twice. In particular, 8 participants were tested both prior to and immediately upon returning from Havana (forming part of both non-exposed and recent test groups), while 1 recently exposed subject was tested again as a remote subject. (See Appendix Figure 3 for a visual breakdown.)

### DEFINITION OF EXPOSURE

“Exposure” here refers strictly to exposure to Havana. The duration of stay in Havana ranged from five to eight months (mean 6.5 months) for recently exposed subjects, and from 1-24 months (mean 21.95 months) for remotely exposed subjects.

For all 26 subjects, we conducted an initial screening across five dimensions: medical history and anthropometric measures, self-reported symptom questionnaires, computerized cognitive assessments, blood tests, and brain imaging (magnetic resonance imaging (MRI) and magnetoencephalography (MEG)). Individuals suspected to have incurred brain injuries (based primarily on self-reported symptom questionnaires) underwent further neurological, visual, auditory, and vestibular assessments, as needed.

### Initial Screening

Medical history and anthropometric measures included travel history, height, weight (to calculate BMI), blood pressure, and heart rate.

Self-rated questionnaires included the Rivermead Post-Concussion Symptoms Questionnaire (RPQ),^3,4^ Migraine Disability Assessment Test (MIDAS),^5^ Headache Impact Test (HIT-6),^6^ Beck Depression (BDI-II) and Anxiety Inventories,^7,8^ Post-Traumatic Stress Disorder Checklist – Civilian (PCL-5),^9^ and Pittsburgh Sleep Quality Index (PSQI).^10^

Cognitive functioning was assessed across the domains of executive functioning, processing speed, attention, working memory, and episodic memory using CANTAB (www.cantab.com). In addition to our core non-exposed group of 12 subjects, cognitive results for our exposed group were compared to 35 healthy age- and sex-matched diplomats not posted in Havana.

Blood testing assessed kidney and liver functions, fasting glucose and insulin levels, lipid panel, complete blood count, thyroid stimulating hormone, and C-reactive protein.

MRI was conducted using a 3-T GE MR750 MRI scanner and included T1, T2, diffusion-weighted, and dynamic contrast-enhanced imaging.

For Diffusion MRI analysis, we used Mrtrix3 software^11^ and Fixel-Based Analysis.^12^ A total of 66 scans were analyzed: 6 from recently exposed subjects, 12 from remotely exposed subjects, 8 from non-exposed subjects, and 40 from healthy age- and sex-matched participants.

Voxel-based blood-brain barrier (BBB) imaging analysis was performed as reported earlier.^13^ The normalized contrast agent accumulation rates were defined as the unit-of-measure for BBB permeability. To calculate overall BBB leakage, the number of voxels with a contrast accumulation rate higher than the 95^th^ percentile of all values in a cohort of control subjects (considered to have a leaky BBB) was divided by total brain volume.^13^ Of the 126 anatomical regions assessed, a brain region was determined leaky when the percent of volume with accumulation rates exceeded three standard deviations of controls. A total of 43 scans were analyzed, including 8 from recently exposed subjects (both before and after exposure), 12 from remotely exposed subjects, and 10 from healthy age- and sex-matched controls.

Resting state MEG data were collected using an Elekta Neuromag whole head 306-channel MEG system.^14^ We used fast Fourier transform (FFT) analysis for studying alterations in brain activity. Periods of paroxysmal slow wave events (PSWEs) were defined as time periods of brain activity in which the median power frequency was less than 6 Hz for more than 5 seconds. Data was analyzed from 11 recently exposed subjects, 15 remotely exposed subjects, and 61 healthy age- and sex-matched controls (21 of whom were recorded in the same MEG, while an additional 40 recordings were obtained from the CamCAN repository (http://www.mrc-cbu.cam.ac.uk/datasets/camcan/).

See Table 1 for a breakdown of clinical and research methods used during Initial Screening. For more details, see Appendix.

**Table 1.**
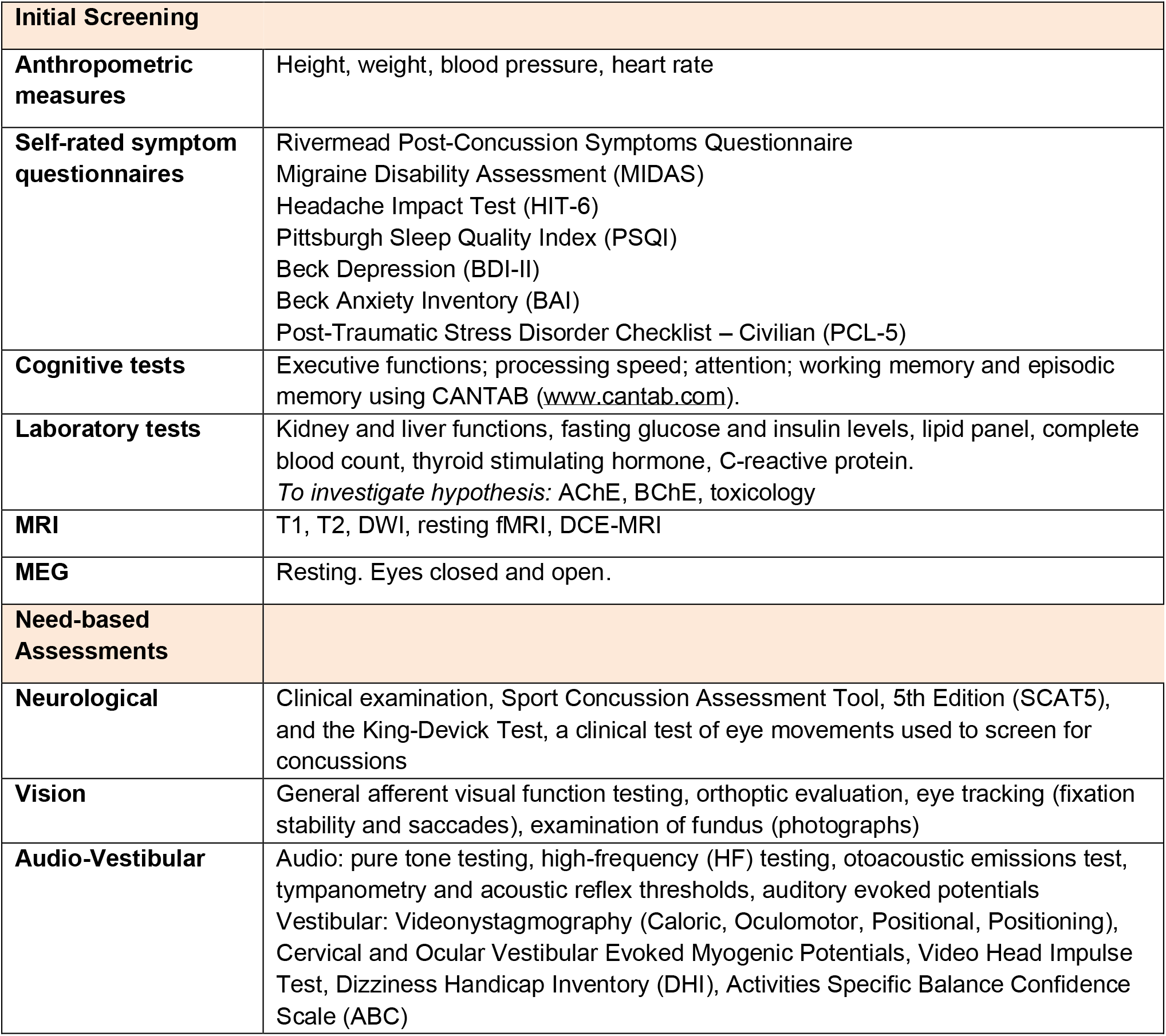
Clinical and Research Measures Used.

### Need-Based Assessments

Individuals who reported symptoms suggesting brain injury underwent further neurological, visual and audio-vestibular assessments.

The neurological assessment included a clinical examination, Sport Concussion Assessment Tool (SCAT5), and the King-Devick Test.

The visual assessment included evaluation for afferent and efferent visual system defects and a detailed assessment of ocular alignment in all positions of gaze as well as horizontal saccadic velocities and fixation stability.

The audio-vestibular assessment included pure tone testing, otoacoustic emissions and immittance testing, tympanometry and acoustic reflex threshold testing, auditory evoked potential testing, a Video Head Impulse Test (vHIT), Videonystamography (VNG), a caloric test, and cervical and ocular vestibular myogenic potential testing.

See Table 1 for a breakdown of clinical and research methods used during Need-Based Assessments. For more details, see Appendix.

### Statistical Analysis

Statistical data analysis was performed using MATLAB and Prism, while Mann-Whitney U and χ2 were used for comparisons, as appropriate. For fibre tractography, statistical analysis was conducted using Connectivity-based Fixel Enhancement (CFE), non-parametric permutation testing using 5000 permutations, and Family-Wise error correction at a P-value of 0.05. Age was included in the analysis as a nuisance covariate. For comparing damage levels between the recently exposed, remotely exposed, and non-exposed groups, a mask was generated for significant fixels, while extracted fibre density values were compared using t-tests. Statistical comparison of BBB leakage in subjects scanned prior to and shortly after exposure were performed using a paired two-tailed Wilcoxon signed rank test. Differences between recently and remotely exposed subjects were assessed using a two-tailed Wilcoxon rank sum test. P-values below 0.05 were considered significant.

## Results

### Initial Screening

#### Medical History

Symptoms included cognitive impairment (concentration and memory), visual impairment (blurred vision and sensitivity to light), audio-vestibular impairment (tinnitus, sensitivity to sound, feeling off balance), and generally reduced well-being (sleep disturbances, fatigue, headaches, irritability) (Table 2).

**Table 2.** Prevalence of Persistent Symptoms.

| Domain | Symptom | No Exposure<br>N=12 (%) | Recent Exposure<br>N=11 (%) | Remote Exposure<br>N=12 (%) | P Value<br>($\chi^2$) |
| --- | --- | --- | --- | --- | --- |
| **Cognitive** | Concentration | 0 | **6 (54)** | **4 (33)** | **0.014** |
|  | Memory | 0 | 3 (27) | **6 (50)** | **0.019** |
| **Dizziness** | Balance | 0 | 3 (27) | **9 (75)** | **0.001** |
|  | Vertigo | 1 (8.3) | 3 (27) | 3 (25) | 0.456 |
|  | Light Headedness | 1 (8.3) | 3 (27) | 3 (25) | 0.456 |
| **Visual** | Blurred Vision | 1 (8.3) | 5 (45) | **6 (50)** | 0.064 |
|  | Light Sensitivity | 0 | 2 (18) | **5 (41)** | **0.038** |
| **Audio-Vestibular** | Tinnitus | 0 | 2 (18) | **8 (66)** | **0.001** |
|  | Sound Sensitivity | 0 | 3 (27) | **8 (66)** | **0.002** |
|  | Vestibular | 0 | 0 | **7 (58)** | **0.000** |
| **General Well-Being** | Sleep | 1 (8.3) | **6 (54)** | **7 (58)** | **0.024** |
|  | Fatigue | 0 | **5 (45)** | **8 (66)** | **0.002** |
|  | Headaches | 0 | **6 (54)** | **11 (91)** | **<0.000** |
|  | Irritability | 0 | 2 (18) | **5 (41)** | **0.0129** |
|  | Nausea | 3 (25) | 3 (27) | **6 (50)** | 0.3652 |

#### Anthropometric measures

Results were similar to what is usually obtained in the Canadian population, with no significant differences between the groups (Appendix Table 2).

#### Blood Tests

Results were similar to what is usually obtained in the Canadian population, with no significant differences between the groups (Appendix Table 3).

**Table 3.** Self-Reported Symptom Questionnaires.

| Domain | Questionnaire | No Exposure<br>N = 12 (%) | Recent Exposure<br>N = 11 (%) | Remote Exposure<br>N = 12 (%) | P value<br>($\chi^2$) |
| --- | --- | --- | --- | --- | --- |
| **Concussion** | RPQ | 2 (16) | 5 (45) | **8 (66)** | **0.0457** |
| **Headaches** | HIT-6 | 0 | 4 (44) N=9 | **7 (58)** | **0.0072** |
| **Sleep** | PSQI | **7 (58)** | **8 (72)** | **8 (72) N=11** | 0.6924 |
| **Migraine** | MIDAS | 0 | 1 (9) | **5 (41)** | **0.0177** |
| **Mental State** | MMS | 0 | 1 (9) | 2 (16) | 0.3443 |
| **Depression** | BDI-II | 0 | 1 (9) | 3 (25) | 0.1396 |
| **Anxiety** | BAI | 0 | 0 (0) | 2 (16) | 0.1310 |
| **PTSD** | PCL-5 | 0 | 1 (9) | 1 (8) | 0.5733 |

#### Self-Reported Symptom Questionnaires

Of the eight questionnaires, three were scored positively by the majority of exposed individuals: RPQ (for post-concussive syndrome), HIT-6 (for headache severity), and MIDAS (for migraine) (Table 3).

#### Cognitive Assessments

We recorded significantly lower performance in spatial working memory within both recently and remotely exposed individuals compared to age-matched controls (P=0.0003). A milder reduction in performance was also found in decision-making quality (Table 4).

**Table 4.** Cognitive Assessment Results.

| TEST | Reaction Time<sup>1</sup> | Paired Associates Learning<sup>2</sup> | Spatial Working Memory<sup>3</sup> | Multitesting Task<sup>4</sup> | Stockings of Cambridge<sup>5</sup> | Cambridge Gambling Task<sup>6</sup> |
| --- | --- | --- | --- | --- | --- | --- |
| **Controls (Mean±SDV, N)** | 371.9±34.6, 45 | 8.9±8.4, 45 | 1.3±1.8, 25 | 2.7±1.9, 32 | 6.0±1.0, 34 | 0.98±0.02, 30 |
| **Recent Exposure (Mean±SDV, N)** | 368.25±20.9, 10 | 8.7±11.16, 10 | 6.9±7.06, 10 | 3.22±2.28, 9 | 5.97±0.85, 8 | 0.97±0.03, 9 |
| **Remote Exposure (Mean±SDV, N)** | 369.9±29.42, 12 | 9.5±7.66, 12 | 9.44±7.8, 9 | 3.78±3.40, 9 | 5.33±0.41, 9 | 0.95±0.05, 11 |
| **P** | 0.6 | 0.9 | **0.0003** | 0.5 | 0.21 | **0.04** |

<sup>1</sup>Median five choice reaction time; <sup>2</sup>Total Errors (adjusted); <sup>3</sup>Number of errors (across all); <sup>4</sup>Number of incorrect responses (total); <sup>5</sup>Mean number of moves; <sup>6</sup>Decision-making quality (total score)

### Need-Based Assessments

#### Neurological Assessment

Results were similar to what is usually obtained in the Canadian population, with no significant differences between the groups.

#### Visual Assessment

Results were similar to what is usually obtained in the Canadian population, with no significant differences between the groups (Appendix Table 4).

#### Audio-Vestibular Assessment

Hearing loss was found in 3 of the 20 exposed subjects tested (15%) but was asymptomatic, and in 2 of the 3, could be attributed to a history of sound exposure prior to exposure. Auditory brain-stem evoked potentials, however, showed long latencies (both absolute and interpeak) in the majority of exposed individuals, with no significant difference between the two exposed groups. Acoustic reflex was also found to be positive in 80% of individuals in both exposed groups (Appendix Table 5). The most consistent finding in the vestibular assessment was the presence of low-threshold, high-amplitude cervical and/or ocular vestibular evoked myogenic potentials. This was found in 33-66% of exposed individuals, with no difference between the exposed groups (Appendix Table 6). Both auditory and vestibular assessments were consistent with brain-stem dysfunction.

### Brain Imaging (MRI & MEG)

#### Clinical MRI

In 4 of the 26 scanned subjects, non-specific white matter hyperintensities in T2 and FLAIR sequences were observed; in one subject, a small capillary telangiectasia in the pons was observed; in another, Chiari I malformation was observed. Extracranial findings in four individuals included mucosal inflammatory changes of the paranasal sinuses. All such findings were considered incidental and similar to what is often seen in healthy individuals, without clinical significance.

#### Diffusion Tensor Imaging (MRI)

A significant difference in white matter integrity between the non-exposed and exposed groups was indicated (P<0.05). Decreased fibre density was observed in the exposed group, predominantly along the right crus of the fornix, past the hippocampal commissure, and projecting into the hippocampus, as well as in the splenium of the corpus callosum (Figure 1).

**Figure 1.**
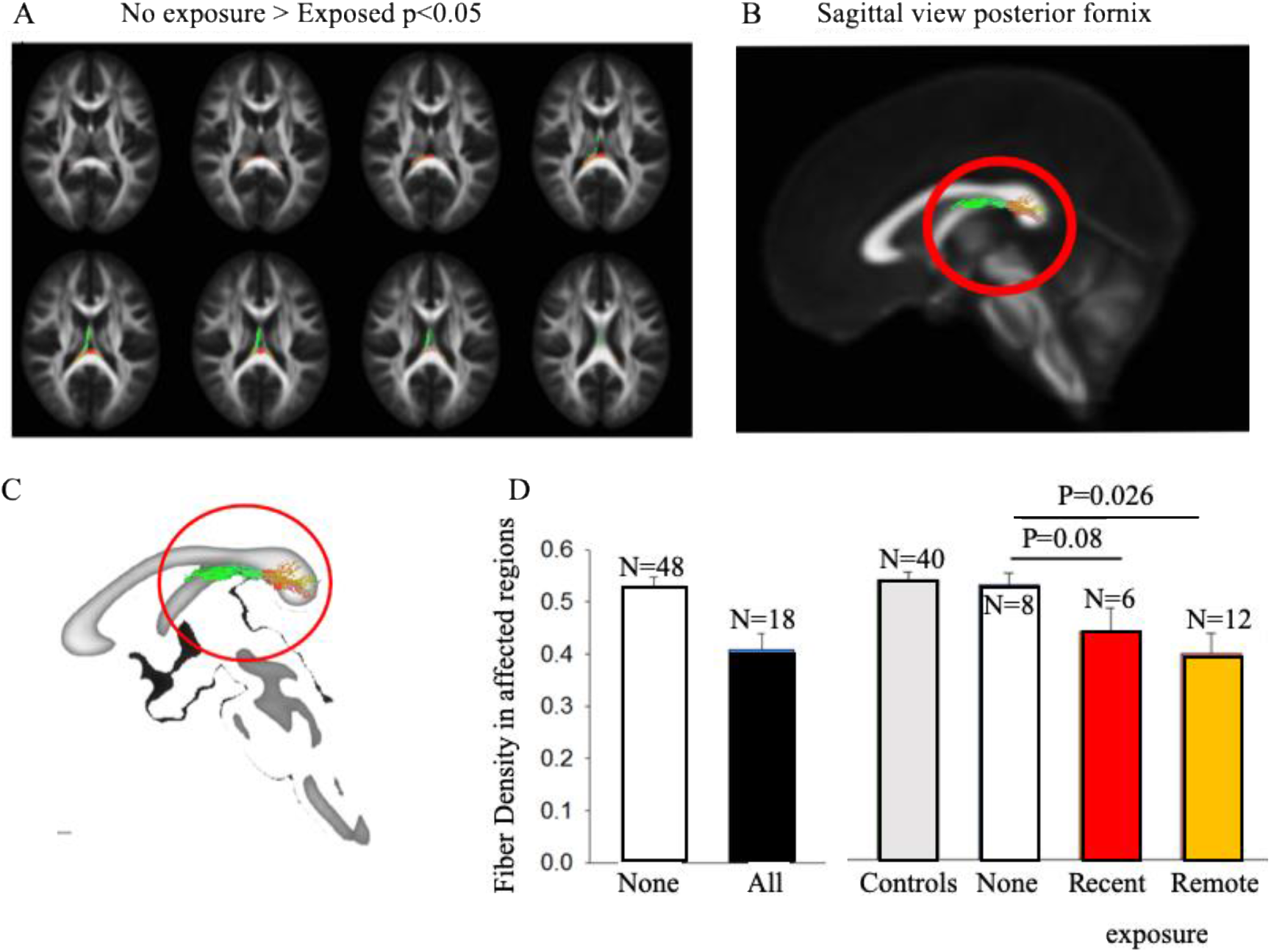
Diffusion Tensor Imaging Reveals Fibre-Specific Changes in Exposed Individuals. A. Streamlines coloured by direction that correspond to fixels indicating a significant difference between the non-exposed and exposed groups (P<0.05, age and error-corrected). B-C. A decrease in fibre density was observed predominantly along the right crus of the fornix, past the hippocampal commissure and projecting into the hippocampus, as well as in the splenium of the corpus callosum. D. Post-hoc analysis indicated decreased fibre density in the remotely exposed group compared to the pre-exposed group (N=8, P=0.026) and all non-exposed control groups (N=40, P<0.001). Recently exposed participants showed a trend for reduced fibre density compared to pre-exposed controls (P=0.08), and significant reduced fibre density when compared to non-exposed controls (P=0.032).

#### Dynamic Contrast-Enhanced MRI

Analysis of 8 subjects tested both before and after living in Havana revealed an increase in the brain volume and number of regions with a leaky blood-brain barrier (BBB). Statistical comparison of the extent of BBB dysfunction in 126 anatomical regions revealed 6 leaky brain regions in post-exposure compared to pre-exposure scans (P<0.05). A leaky BBB was mainly found in the right hemisphere and included the right basal forebrain, anterior insula, posterior orbital, superior frontal, and superior occipital gyri (Figure 2).

**Figure 2.**
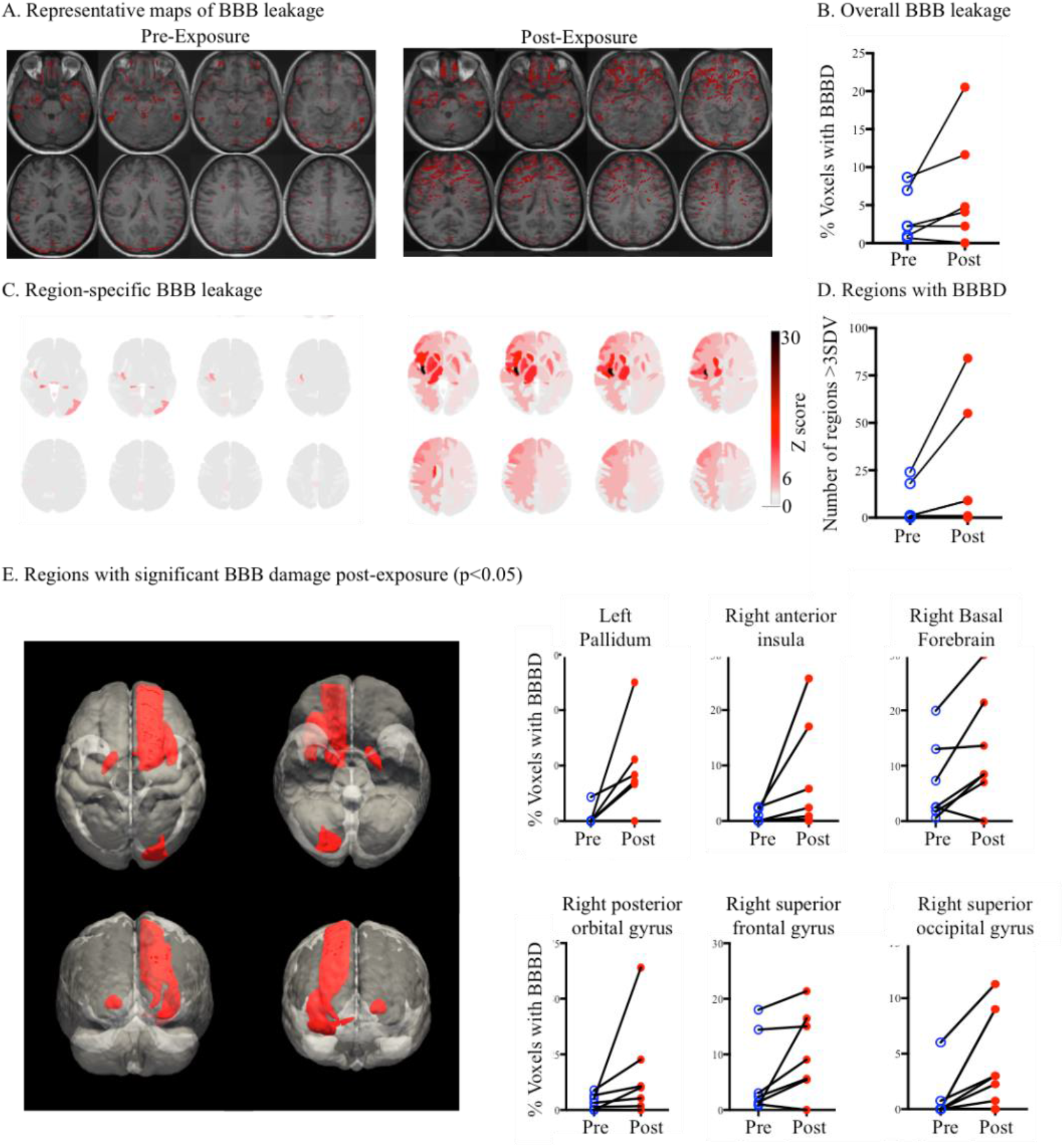
Blood-Brain Barrier (BBB) Dysfunction Following Exposure. A. A typical scan from an individual prior to living in Havana for approximately 6 months as well as after. Voxels with a leaky BBB (>95 percentile of controls) are marked in red. B. Residence in Havana was associated with an increase in the brain volume and number of regions with a leaky BBB (Wilcoxon, P=0.06). C. Regional analysis showed brain regions with a leaky BBB > 3 standard deviations of controls, mainly in the right hemisphere. D. The number of leaky regions pre- and post-exposure (Wilcoxon, P=0.06). E. 6 (out of 126) brain regions were found to show a statistically significant increase in contrast accumulation (i.e., a leaky BBB) in post-compared to pre-exposure scans (P<0.05).

Brain regions with a high contrast agent rate of accumulation (i.e., a leaky BBB) were similar in both exposed groups, although to a slightly lesser extent in the remotely exposed group. Similar brain regions, specifically the right basal forebrain, anterior insula, and superior frontal gyrus, were found to be leakier in both exposed groups compared to non-exposed controls (Figure 3).

**Figure 3.**
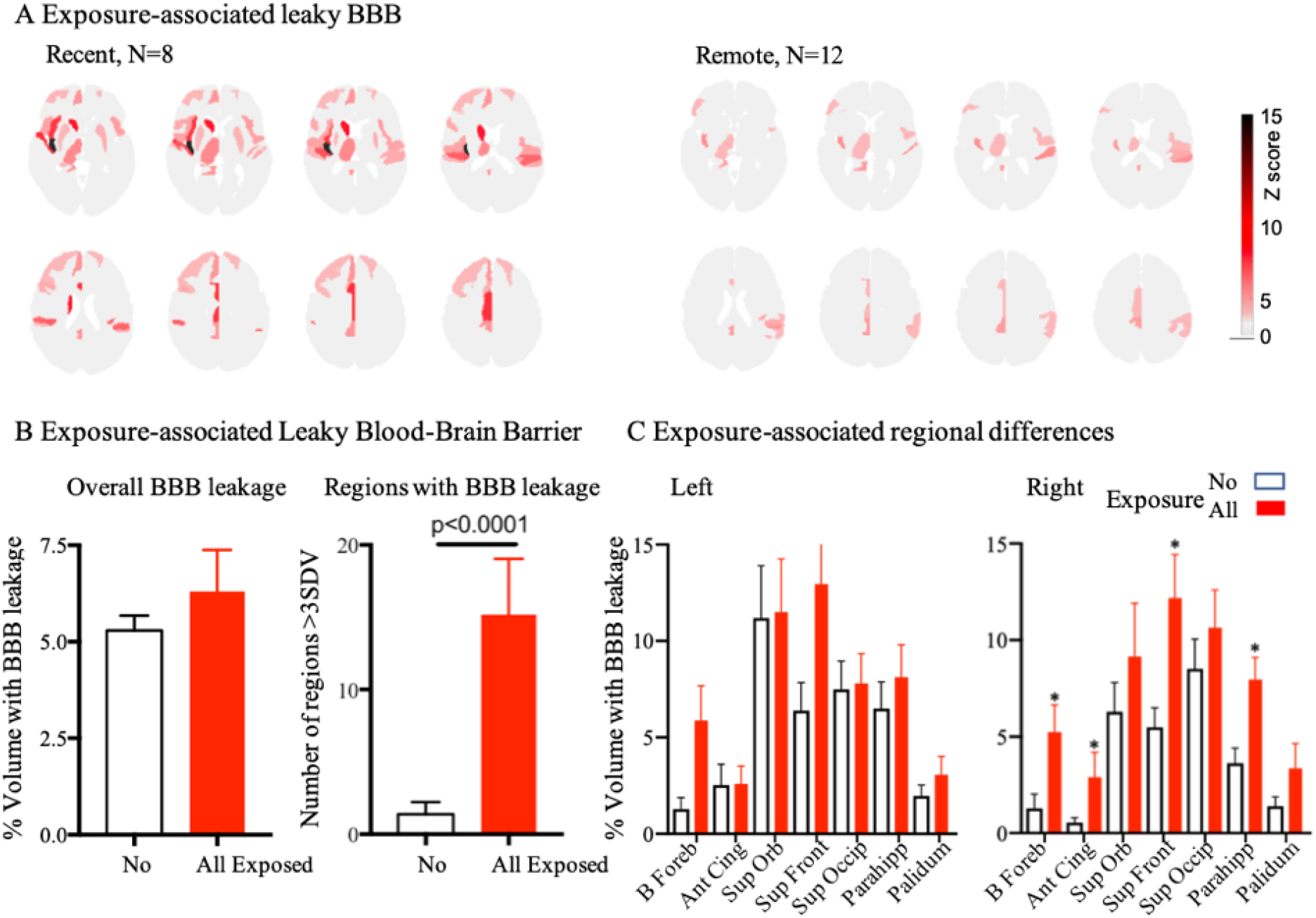
Region-Specific Blood-Brain Barrier (BBB) Dysfunction Following Exposure. A. Averaged Z-score for each brain region in recently (left) and remotely (right) exposed groups. Note the similarity in regional BBB pathology, with a more pronounced dysfunction in the recently exposed group. B. Overall percentage of brain volume (left graph) and number of regions (right graph) with a leaky BBB, in all exposed compared to non-exposed controls. C. Regions found to be leaky after exposure compared to before exposure (n=8, Figure 2), were next compared between the entire exposed (N=25) compared to the non-exposed groups. We also tested the hypothesis that BBB integrity would be impaired in the parahippocampus gyrus due to its well-recognized role in spatial memory.^32^ Indeed, the right parahippocampal cortex (but not the left) displayed a leaky BBB. Regions: Basal forebrain (B Foreb), Anterior Cingulate (Ant Ching), Superior Orbital (Sup Orn), Superior Frontal (Sup Fron), Superior occipital (Sup Occip) and parahippocampal (Parahipp) gyri.

#### Magnetoencephalography (MEG)

Exposed individuals showed an increase in power in the delta frequency range with decreased alpha. These changes were due to a transient, intermittent slowing of brain activity, termed “paroxysmal slow-wave events” (PSWEs). PSWEs were rarely detected in healthy controls, including test subjects prior to exposure. By contrast, PSWEs were significantly more common in both exposed groups and were diffusely distributed in both hemispheres, more prominently on the right (Figure 4).

**Figure 4.**
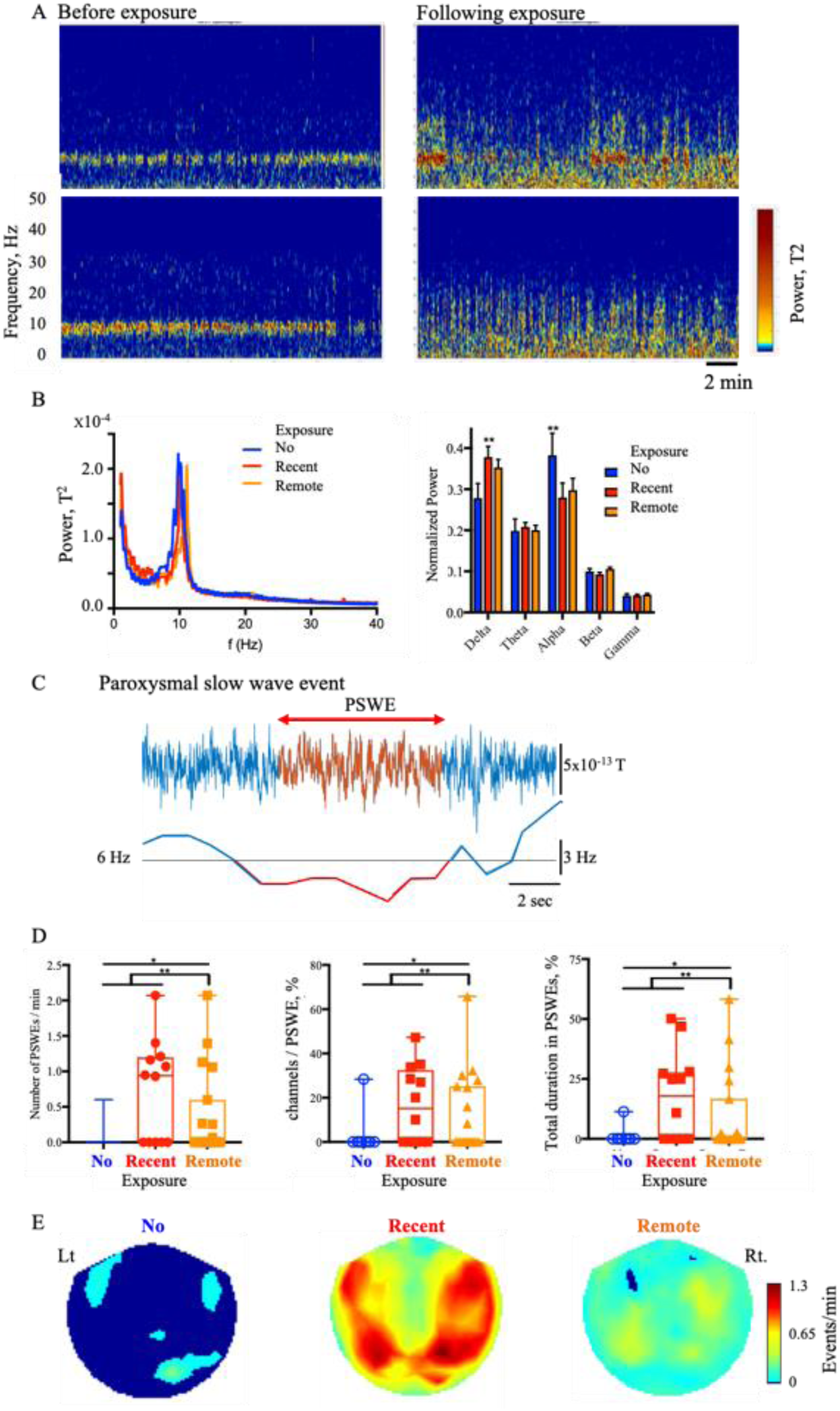
Magnetoencephalography Reveals Paroxysmal Slowing of Brain Activity. A. Spectrogram with eyes closed from two recently exposed individuals pre- (left) and post- (right) exposure. Note the clear dominant 10Hz rhythm prior to exposure and intermittent slowing following. B. Group comparison shows significantly lower power in the alpha and higher in the delta band in both recently and remotely exposed groups. C. Slowing of activity was composed of transient, paroxysmal slow wave events (PSWEs). The upper trace is the original MEG recording, and the calculated median frequency below. A PSWE was defined as the time window in which median frequency was < 6 Hz for > 5 sec. D. Graphs showing the number of PSWEs/minute (left), % of MEG channels with PSWEs (middle) and % of recording time in which the individuals spent in PSWEs (right). E. Scalp mapping showing spatial distribution of PSWEs.

### Key Finding & Emergent Hypothesis: Exposure to Cholinesterase Inhibitors

Our brain imaging findings—in particular the consistent involvement of the basal forebrain, a key cholinergic nucleus—raised the hypothesis that one or more environmental neurotoxins targeting the brain’s cholinergic system may underlie the observed injury.

To support our hypothesis, serum samples tested for acetyl- and butyryl-cholinesterase activity showed reduced enzyme activity in individuals recently returned from Havana compared to remotely exposed and control groups (Figure 5).

**Figure 5.**
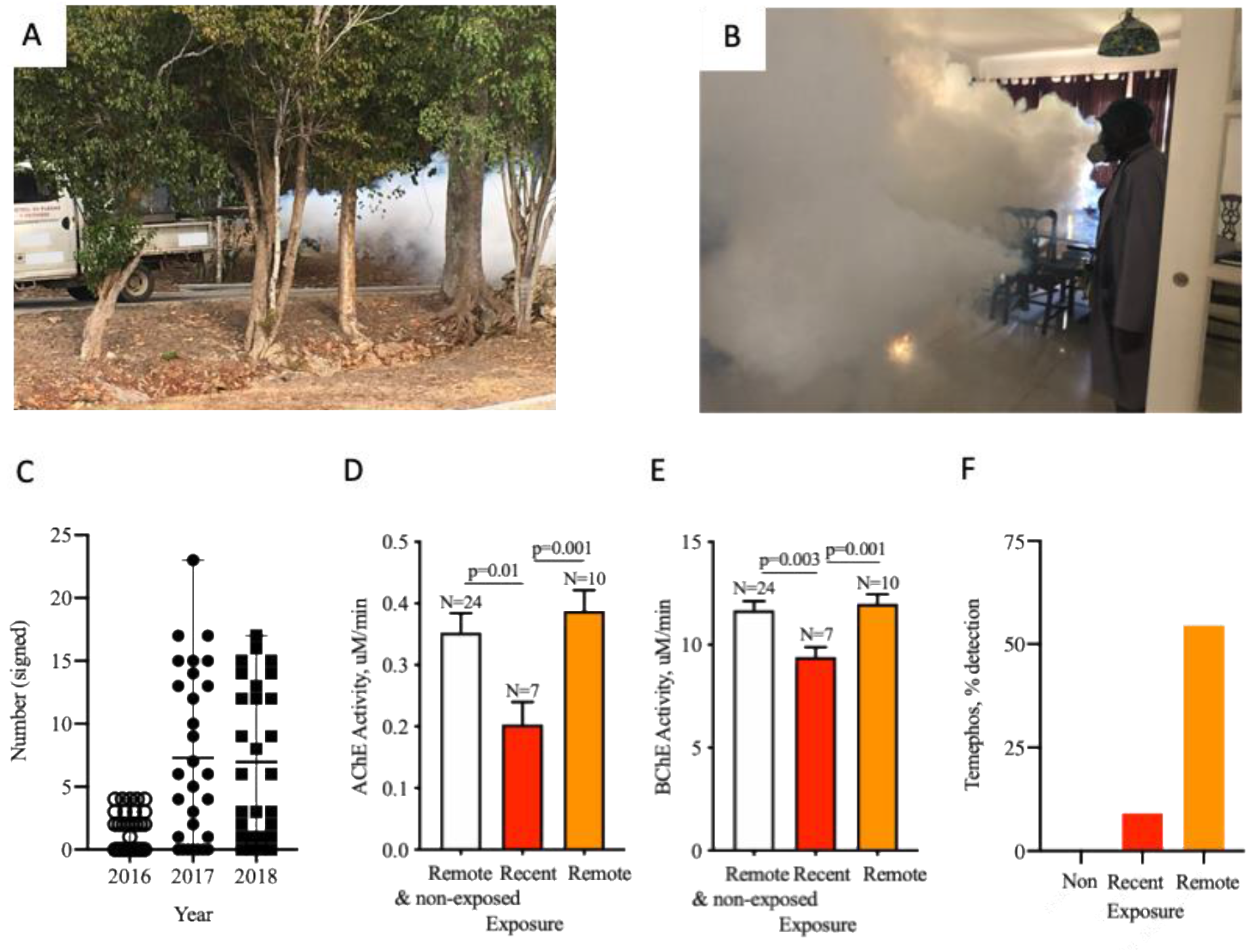
Fumigation in Havana and Cholinesterase Inhibitor Exposure. A-B. Outdoor and indoor fumigation efforts in Cuba for the eradication of mosquitos as initiated by the Cuban government (see also ^16,17^). C. Number of authorized annual outdoor fumigations in Canadian diplomatic residences in Cuba between 2016-2018. Note the sharp increase in frequency during 2017, immediately prior to the appearance of symptoms (mean and range). D-E. Both acetyl- and butyryl-cholinesterase activities in serum were reduced in individuals recently exposed. F. Percentage of detection of the organophosphate metabolite Temephos in exposed individuals (see Appendix Table 7).

Mass-spectrometry furthermore confirmed the presence of cholinesterase-inhibiting insecticides, including Temephos, an organophosphorus used in Cuba against mosquito larvae,^15^ and 3-phenoxybenzoic acid (3-PBA), a common pyrethroid (insecticidal) metabolite. Temephos was detected in six of ten remotely exposed individuals, compared to one recently exposed individual and none of the controls (P<0.001). 3-PBA was found in the majority (62%) of exposed individuals, with no significant difference between the two groups (Appendix Table 7).

## Discussion

Using a multimodal, quantitative, and control-tested approach, our results confirm brain injury, specify the regions involved, and suggest a likely etiology underlying the Havana Syndrome in the form of environmental exposure to neurotoxins affecting the brain’s cholinergic system. We more narrowly suggest organophosphorus insecticides as a likely source.

Though other sources of neurotoxins are possible, our insecticidal hypothesis gains contextual support given Cuba’s well-documented efforts to aggressively mitigate the spread of the Zika virus by means of mass indoor and outdoor fumigations in 2016 and thereafter.^16,17^ Canadian Embassy records furthermore confirmed a significant increase in the frequency of fumigations around and within staff houses beginning January 2017, concurrent with reported symptoms (Figure 5C).

Whether in its broad or narrow form (cholinesterase-inhibiting neurotoxins in general or organophosphorus insecticides in particular), our hypothesis provides a plausible alternative to the theory of “unknown energy exposures”^1^ that have dominated media reports, and as such carries potential public health and clinical ramifications.^18,19^

### Public Health

If our narrow hypothesis of insecticide-induced neurotoxicity is correct, our findings suggest a course of action that affects not only the physical health of diplomats and their family members that are or will be stationed in Havana, but the health of the local Cuban population (where heavy fumigation continues) and of all other populations where such insecticides are heavily used.

Next steps may include collaborating with Cuban scientists (and elsewhere) to test for neural injury among the local population; testing American diplomats previously stationed in Havana for the presence of Temephos and/or other metabolites in the blood; monitoring exposure levels and serum activity; urging the Cuban government to limit the use of organophosphorus insecticides or to use non-organophosphorus insecticides of equivalent efficacy to protect humans from neurotoxic damage while continuing to control the spread of Zika; and lobbying for the ban of organophosphorus insecticides worldwide, with recommendations for alternative forms of pest control. Further research is also required to better understand genetic and epigenetic mechanisms underlying individual differences in sensitivity to organophosphorus insecticides.^20,21^

### Clinical and Diagnostic Methods

We demonstrate the effectiveness of a multimodal approach for diagnosing atypical brain injuries and suggest new diagnostic modalities to quantitatively assess changes in brain structure and function.

Though such diagnostics are primarily intended for research, we nevertheless propose that clinical researcher-practitioners consider these novel techniques valuable diagnostic biomarkers when presented not only with such difficult-to-diagnose conditions as the Havana Syndrome, but also in more common neurological illnesses, including traumatic and degenerative brain disorders.^13,22^ Previous studies of the Havana Syndrome demonstrated the difficulty of diagnosing concussions and other presentations of brain injury using available clinical tools, while the use of research tools and quantitative analysis in the present study (including fibre tracking, BBB imaging, and MEG) allowed the identification of specific brain networks and patterns of injury in our cohort. Though these are merely research tools, we demonstrate their potential in aiding the clinical diagnosis of brain disorders.

### Biology of Disease

While acute exposure to cholinesterase inhibitors causes a typical clinical presentation that is usually easy to recognize (constriction of pupils, salivation, loss of bowel and bladder control, seizures, and reduced consciousness), long-term or repeated exposure to low levels of cholinesterase inhibitors are more difficult to diagnose and may not show any acute symptoms.

Though the neurotoxic effects of low-dose exposure to cholinesterase inhibitors have been suggested in other contexts, our study specifies how the brain is affected and suggests how injury to specific cholinergic centres can underlie a profound symptomatology, including changes in brain stem functions, white matter fibre integrity, and a corresponding reduction in spatial memory (for a full breakdown, see Appendix).

Epidemiologically, our study broadens the scope of the problem of exposure to cholinesterase inhibitors beyond agricultural pesticide use.

### Alternative Hypotheses

Despite prevailing theories of acoustic attacks in the media, the highly focused and consistent pattern of injury among exposed individuals is difficult to explain if resulting from energy exposures. There were furthermore no acute incidents in most individuals; symptoms developed gradually over the course of their stay in Havana.

Though insecticides are a likely source of organophosphates in the present cases, others cannot be definitively ruled out. Organophosphorus warfare agents are both common and available, and historically, several nerve agents of the G-(tabun (GA), sarin (GB), chlorosarin (GC) and soman (GD)) and V-series (VE, VR, VS and VX) have been deployed not only in warfare but also in acts of terrorism^23^ and high-profile assassinations.^24^ While in these previously reported incidents high-dose exposure led to acute poisoning, low-dose exposure may not be associated with typical acute symptoms.^25^ The brain changes we found are also consistent with the reduced volume of the right insular and temporal cortices reported in victims of the 1995 sarin attack in Tokyo.^26^

Though most subjects in our study did not report an acute event of pressure or sound in contrast to previous reports of American diplomats residing in Cuba,^1,2^ symptoms were similar to those reported by them,^2^ including headaches and difficulties in concentration, balance, vision, and sleep. The shared symptoms, location, period, and relative duration of time all point to a shared etiology.

### Further Evidential Support

Acute poisoning with organophosphates has previously been shown to result in a robust opening of the blood-brain barrier,^27^ while injury to the basal forebrain has previously been shown (in animal studies) following acute or chronic exposure to organophosphates.^28^ Slowing of brain activity, although not a specific finding, has also been previously reported to follow exposure to organophosphates.^29^

The most direct evidence of the effects of low-dose exposure to organophosphates was published by Bowers and colleagues (1964),^25^ who analyzed the clinical signs presented by 96 young male volunteers following percutaneous exposure to a low dose of an organophosphate compound (likely VX). No signs of acute toxicity were presented, but a “state of altered awareness” characterized by difficulty sustaining attention and the slowing of intellectual and motor processes, in addition to feelings of agitation, anxiety, and confusion, was reported.^25^ Tinnitus, vestibular, cognitive, psychomotor dysfunction, attention deficits, and memory impairment have all been reported following low-dose cholinesterase inhibitor exposure in humans.^30,31^

For further details regarding brain injury by cholinesterase inhibitors, see Appendix.

### Recovery

Blood-brain barrier leakage and abnormal MEG activity were less pronounced in remotely exposed individuals compared to the recently exposed group; this suggests that injury may be at least partially reversible, and that recovery may be expected once exposure is halted. On the other hand, DTI analysis for fibre integrity suggested a slightly more pronounced reduction in remotely exposed individuals and may indicate progressive axonal injury. Follow-up is required to better understand the natural course of brain injury in affected individuals.

## Conclusion

We report clinical, imaging, biochemical, and toxicological evidence consistent with the hypothesis of overexposure to cholinesterase inhibitors as the cause of brain injury in our cohort. While other causes cannot be ruled out, our findings point to an environmental (and possibly insecticidal) risk with immediate implications for prevention, screening, and follow-up of individuals in the context of exposure to such neurotoxins.

This is the first brain imaging study documenting blood-brain barrier pathology in patients tested both before and after acquired brain injury.

## Data Availability

All data is included

## Acknowledgements

This research was funded by Global Affairs Canada. We would like to thank the Canadian diplomats and their families in participating in this study, as well as Kay Murphy for technical assistance, Eli Burnstein for editing the manuscript, and Luc Palombo with the illustration of Appendix Figure 2.

MEG control data collection and sharing for this project was provided by the Cambridge Centre for Ageing and Neuroscience (CamCAN). CamCAN funding was provided by the UK Biotechnology and Biological Sciences Research Council (grant number BB/H008217/1), together with support from the UK Medical Research Council and University of Cambridge, UK.

## Appendix

### The Cholinergic System and Brain Regions Involved

#### Nucleus Basalis and Basal Forebrain

The nucleus basalis within the basal forebrain includes cholinergic neurons with widespread connections to other parts of the brain, and which are thought to be involved in the modulation of neuronal activity during sleep-wake cycle and attention. Injury to basal forebrain cholinergic neurons may thus explain related symptoms reported in our cohort as well as periods of slowing in brain activity found in MEG (see Figure 5 in main article above). The diffuse pattern of slowing observed, while predominantly in the right hemisphere, supports a diffuse change in brain function as can be induced directly by toxins or indirectly by affecting the basal forebrain’s cholinergic fibres, in contrast to focal injury more characteristic of a traumatic event.

#### Brainstem Nuclei

In addition to the basal forebrain, brainstem nuclei are the other source of brain cholinergic projections. Evidence for brainstem injury was found on audio-vestibular tests (including positive acoustic reflex and vestibular-evoked myogenic potentials, as well as delayed latency in brainstem evoked potentials in the majority of exposed individuals). These results are consistent with previous reports on auditory dysfunction and delayed brain stem evoked potentials following chronic exposure to agricultural organophosphates.^33,34^ The vestibular system, through cholinergic fibres, also alters hippocampal rhythms and is involved in spatial memory, observed to be affected in our cohort.^35^

#### Pedunculopontine Tegmental Nucleus

The other principal cholinergic nucleus is the brainstem pedunculopontine tegmental nucleus (PPN), which is involved in mechanisms of arousal and behavioral state control and participates in control of locomotion and muscle tone. Malfunction of the PPN has a role in gait impairment and balance,^36^ and is consistent with the abnormal processing of auditory input found in the majority of exposed individuals (see Appendix Tables 7-8 below).

#### Cholinesterase Inhibitors and The Blood-Brain Barrier

We found microvascular damage (a leaky BBB) in most exposed individuals, with a significant region-specificity when comparing individuals tested both pre- and post-exposure and in all exposed compared to non-exposed controls. While data on the effects of low-dose exposure is scarce, acute poisoning with organophosphates has been shown to result in robust BBB opening.^27,37^ In addition, several studies have demonstrated that organophosphorus pesticides have a direct effect on microvasculature and cause increased permeability in in-vitro models of the BBB.^37^ Particularly interesting was the region-specificity involved. Out of 126 brain regions tested, the right basal forebrain and anterior insula significantly and consistently showed microvascular damage in both exposed groups compared to controls. The basal forebrain is a key brain cholinergic nucleus that has a key role in arousal, attention, memory, and the sleep-wake cycle. The right anterior insula is considered to be associated with sensory processing and body awareness.^38^The basal forebrain sends input to the hippocampus and parahippocampal gyrus via the fornix, which was found to show reduced fibre density in the exposed group. Interestingly, the right parahippocampal gyrus has been shown to be critical in spatial memory processes,^32^ which were impaired in our cohort (see Appendix Table 5 below). The reason for the dominancy of the right brain in the observed injury is not known.

### Methods

#### Neurological Assessment

The neurological assessment included a full clinical examination, Sport Concussion Assessment Tool - 5th Edition (SCAT5),^39^ and the King-Devick Test, a clinical test of eye movements used to screen for concussions.^40^

#### Visual Assessment

Subjects were evaluated for afferent and efferent visual system defects including infranuclear pathways for cranial nerves 2,3,4 and 6. All testing was conducted within the Pediatric Ophthalmology and Strabismus unit at the IWK Health Centre. Each evaluation included a detailed history to determine the presence and nature of any reported visual symptom.

##### General visual assessment

The afferent visual system was evaluated by visual acuity testing (near and distance), contrast sensitivity function, and a full evaluation of pupils. Refractive status was assessed but formal cycloplegic refractions were not conducted. Un-dilated fundus photographs were obtained and reviewed by an ophthalmologist. Additional tests were done in the presence of specific signs or symptoms. This included visual electrophysiology, accommodative function, color vision, exophthalmometry, intra-ocular pressure (I-Care device not requiring anesthetic) and reassessment of visual function with appropriate lenses in place in situations of uncorrected/mis-corrected refractive errors or presbyopia.

##### Orthoptic evaluation (emphasis on efferent visual system)

This consisted of a detailed assessment of ocular alignment in all positions of gaze using standardized clinical tests; gross assessment of ocular movements with attention to the extent of excursions; and integrity of eye movement sub-systems (smooth pursuit, saccades, and vergences). Special attention was directed to convergence ability when nearby.

##### Orthoptic evaluation (emphasis on binocular vision)

Testing involved the assessment of stereo-acuity and fusional amplitudes (nearby and at a distance) using standardized clinical tests.

##### Eye movement recordings

The Eyelink 1000 eye tracking system was used to assess horizontal saccadic velocities between antagonist extra-ocular muscles and fixation stability on at central near fixation target.

#### Audio-Vestibular Assessment

Our audio-vestibular assessment was conducted according to clinical guidelines on 20 of our subjects (10 recently exposed, 10 remotely exposed).

The assessment included pure tone testing (according to the American National Standards Institute (ANSI) standards and high-frequency (HF) testing (at 9 kHz, 10 kHz, 11.2 kHz, 12.5 kHz, 14 kHz, 16 kHz and 18 kHz).

Audiometry was performed with a Madsen Astera (GN Otometrics, Germany) audiometer with Sennheiser HDA-200 supra-aural headphones. Otoacoustic emissions test and Immittance Testing.

Tympanometry and acoustic reflex thresholds were performed with a diagnostic impedance meter *Zodiac* (Madsen, Natus). For evaluating acoustic reflex thresholds, ≥100 dB HL were considered increased. An absent reflex was observed when there was no measurable change in immittance (0.02mm) at 105 dB. Auditory evoked testing was conducted with click stimulus at 80 dB HL, monaural, 11.7 clicks per second, 2000 sweeps, alternating polarity (High pass filter = 100 Hz, low pass = 3000 Hz).

Our vestibular assessment included a Video Head Impulse Test (vHIT, Micromedical Spectrum 9.1 Visual Eyes System); Videonystamography (VNG): Oculomotor (gaze, saccades, tracking, optokinetics), positional, positioning, and caloric testing (Micromedical Spectrum 9.1). Cervical (sternocleidomastoid) and ocular vestibular myogenic potentials were obtained using a 500-Hz, 95-dB nHL tone burst, with a 2-cycle rise time, 1-cycle plateau, and 2-cycle fall time (Biologic Navigator Pro).

#### Magnetic Resonance Imaging (MRI) Analysis

Diffusion-weighted MRIs were acquired with an Oblique-Axial SS-EPI sequence with magnetic gradients with b-values of 1000 s/mm^2^, 60 directions, and the following specifications: FOV 22cm, 77 axial slices, voxel size 2 × 2 × 2 mm^3^, TR = 8s, and an acceleration factor of 2. The image acquisition sequence also included 7 b=0 interleaved acquisitions to facilitate motion correction. To facilitate image distortion correction, an additional scan of 7 b=0 images having phase encode blip polarity reversal was acquired. DCE-MRI was conducted according to published protocols (GE Discovery MR750, 3T, FOV 24cm, slice thickness 6mm, 192×192 matrix, flip angle 15°, TR/TE 4.1/2.1ms) before and after intravenous injection with the magnetic contrast-agent gadoteridol (0.1 mmol/kg, ProHance, Bracco Imaging Canada, Montreal, QC). DCE-MRI (N=40) was performed on 10 recently exposed and 12 remotely exposed individuals and compared to 16 non-exposed controls (including a previous cohort). Of the 7 individuals scanned twice, 6 were scanned both immediately prior to leaving for Havana and a few (1-6 days) after returning, while 1 individual (who was posted for 13 months in Havana) was scanned 2 days after returning to Canada as well as 2 months later.

#### Diffusion MRI Analysis

We used MRtrix3 software^11^ and Fixel-Based Analysis (FBA).^12^ Preprocessing of diffusion images (N=65) included denoising, removal of Gibbs ringing artefacts, and correction of distortions induced by susceptibility off-resonance fields, eddy currents, and head motion.^41^ In addition, we used bias field correction and global intensity normalization across the normative healthy participants using group-wise registration to a study-specific fractional anisotropy (FA) template. Constrained spherical deconvolution was used to obtain single-fibre white matter response functions and averaged across all participants. Obtained response function was used to compute Fibre Orientation Distribution (FOD) images following the up-sampling of diffusion images to voxel size of 1.3 × 1.3 × 1.3 mm.^3^ A study-specific and unbiased FOD template was generated using all normative healthy participants (N=40).

Participants were incorporated to the cohort by correcting for the bias field and performing global intensity normalization using the FA template generated. FOD images for all participants were then registered to the created template. Registrations were used to warp subject masks to the template and obtain an intersection template mask subsequently used for the segmentation of the FOD template into a fixel template mask.

Segmentation involved thresholding the peak amplitude of positive FOD lobes at 0.3 to constrain the fitting of fixels to white matter fibre populations and mapping obtained fixels to a fixel mask. Next, subject FOD images were segmented into fixels and values for Fibre Density (FD) were computed per fixel for each subject following spatial transformation of the FOD image to the template space.

Spatial and orientational matching between subject and template fixels involved reorientation of all subject fixels using FOD registration warps and a correspondence process to assign each template fixel to a matching subject fixel, across all subjects. Using the template fixels and the subject-to-template Jacobian matrix warps, Fibre Cross-section (FC) was obtained for each fixel across subjects and then normally distributed by performing a log transformation of the metric. A conjunction of the two metrics, Fibre Density and Cross-section (FDC), was then computed by multiplying the two metrics.

Whole-brain probabilistic tractography was performed on the FOD template using an FOD amplitude cutoff of 0.1 to generate 20 million streamlines. The tractogram was then filtered using Spherical-deconvolution Informed Filtering of Tractograms (SIFT) to 2 million streamlines to reduce biases from the tractography algorithm. Group statistical comparisons were performed independently on FD, FC, and FDC for every fixel using a General Linear Model to investigate microstructural differences in white matter tracts between the unexposed cohort and the exposed cohort.

#### Voxel-Based BBB Imaging Analysis

Analysis for BBB integrity was performed as published.^13,42,43^ Briefly, pre-processing included image registration and normalization to MNI coordinates using SPM12 (University College London, www.fil.ion.ucl.ac.uk/spm). The accumulation rate of the contrast-agent during the slow enhancement period of the scan (6-20 mins.) was calculated for each voxel. To compensate for physiological (e.g., heart rate, blood flow) and technical (e.g., injection rate) variabilities between scans, each voxel’s accumulation rate was normalized to that of the superior sagittal sinus. The normalized contrast-agent accumulation rates were defined as the unit-of-measure for BBB permeability, with near-zero/negative values reflecting BBB-protected tissue and positive values representing tissue with tracer accumulation due to cross BBB extravasation. Abnormally high BBB permeability was defined using an intensity threshold of the 95th percentile of all values in a cohort of control subjects.^13^

#### Region-Specific BBB Imaging Analysis

To quantify region-specific BBB leakage, each scan was segmented into 126 anatomically/functionally significant areas in accordance with the MNI brain atlas (https://github.com/neurodebian/spm12/tree/master/tpm). The number of voxels with abnormally high BBB permeability (contrast accumulation rates exceeding the above-mentioned intensity threshold^13^) was quantified within each region and divided by the total number of voxels comprising the region. This ratio was defined as the percent of region volume affected by abnormally high BBB permeability, and was used as the measure of region-specific BBB damage.

#### Magnetoencephalography (MEG)

Resting state MEG data were collected using an Elekta Neuromag whole head 306-channel MEG system.^14^ Standard operating procedures were utilized for continuous head position monitoring. Electrooculogram (EOG) and electrocardiogram (ECG) data were recorded for the removal of physiological artifacts. Research participants were scanned in the seated position in a magnetically shielded room with active shielding enabled. Two resting-state datasets were acquired: one with the subject’s eyes open and the other with eyes closed, each with a 15-minute duration. MEG data were sampled continuously during all resting state recordings at 1000 Hz and band-pass filtered at 0.1-330 Hz.

#### MEG Data Analysis

Data was analyzed from 11 recently exposed and 12 remotely exposed individuals and compared to 61 age-matched controls. Controls included 11 non-exposed individuals from the present cohort and 10 healthy controls recorded in the same MEG from a previous cohort. Additional data from 40 age-matched healthy controls (mean age=42.6 ± 11.15 years) was obtained from the CamCAN repository (available at http://www.mrc-cbu.cam.ac.uk/datasets/camcan/). Preprocessing using signal space separation^44^ was conducted to account for active shielding and to remove sources external to the MEG helmet. Additional analysis was performed on 10-15 min eye-closed recordings using in-house Matlab scripts: Data was down-sampled to 250 Hz and band-pass filtered in the range 1.0-50 Hz.

Independent component analysis (ICA) was used to identify components correlated with EOG and ECG time courses due to vertical eye movements and blinks, as well as cardiac signals. The corresponding components were removed, and the data was visually inspected in the time domain to remove time segments containing clear artifacts. Spectral analysis by fast furrier transform (FFT) and bandwidth power (1-3, 3-8, 8-12, 12-20, and 20-50 Hz) were calculated for each channel buffered into 2-second long epochs with 1-second overlap, averaged across all epochs, normalized to the total power of the respective channel, and averaged across all channels to compare between subjects. For each activity epoch, the median-power frequency (MPF) was extracted. Periods of paroxysmal slow wave events (PSWEs) were defined as time periods of brain activity in which the MPF was < 6 Hz for > 5 seconds.

#### Serum Cholinesterase Activity Assays

Serum acetyl-cholinesterase (AChE) and butyryl-cholinesterase (BChE) activity were measured in a microtiter plate assay using a SpectraMax M3 according to published methods.^45^ 10μl of 1:40 diluted serum in Tris-HCl buffer was incubated in tetraisopropyl pyrophosphoramide (iso-OMPA, 100μM), a specific BChE inhibitor, or 1,5-bis(4-allyldimethylammoniumphenyl) (BW, 20μM), a specific AChE inhibitor, for 20 minutes. Following incubation, diluted serum was added in triplicates to a 96-well plate. Readings of AChE and BChE activity were repeated at 2-minute intervals for 20 minutes at 405nm following the addition of acetylthiocholine iodide (ATCh, 1mM) or s-butyrylthiocholine iodide crystalline (BTCh, 10mM) substrates, respectively, and Ellman’s reagent (10mM). A cysteine standard of known concentrations was used to calculate enzyme activity.

#### Sample Preparation and Analysis for Toxicology

Serum for mass spectrometry analysis samples were prepared as described elsewhere.^46^ QuEChERS (Quick Easy Cheap Effective Rugged Safe) salt mixtures (50 mg MgSO_4_ and 12.5 mg NaCl) was added to the 125 µL of serum sample in 1.5 mL microcentrifuge tube to facilitate phase separation and extraction of target analytes. Samples were vortexed for 15 seconds to break up salt agglomerates. 125 µL of acetonitrile (ACN) was added to the samples to extract target analytes. The samples were shaken vigorously by hand for 1 minute. Thermo Scientific Accela UHPLC system with the CTC Analytics PAL autosampler coupled to the Exactive Plus orbital ion trap mass spectrometer was used for all sample analyses.

A total of six insecticides or their metabolites reported to be used in Cuba and neighboring countries were chosen as targets, including Cypermethrin, a pyrethroid insecticide, two active ingredients of serpol: Tilt (propiconazole - triazole fungicide) and permethrin (a pyrethroid insecticide), as well as deltamethrin chlorpyrifos and Temephos—both organophosphorus insecticides. The final target analyte chosen for this study was 3-Phenoxybenzoic acid (3-PBA), a common pyrethroid metabolite.

#### Cholinesterase Inhibitors and Insecticides

Cholinesterase (ChE) is one of the key enzymes required for the proper functioning of the nervous systems of humans, other vertebrates, and insects, breaking down the neurotransmitter acetylcholine and preventing its action in the junction between nerves and muscles as well as between two nerve cells.

While the source of exposure to toxins of the cholinesterase inhibitor family is not yet confirmed in our study, the use of insecticides readily and evidentially suggests itself. Importantly, certain chemical classes of pesticides, such as organophosphates and carbamates, work against insects by inhibiting the action of cholinesterase, but can also be poisonous to humans.

Most of the literature on low-level exposure to cholinesterase inhibitors (ChEIs) in humans is epidemiological in nature, resulting from dietary ingestion and occupational exposure.^47^ The major concerns related to such exposure are delayed effects on the function of the nervous system and increased prevalence of cognitive, behavioral, and psychomotor dysfunction.^30,48,49^ The exact mechanisms of damage, however, are not known, and additional targets for these toxins (in addition to ChE inhibition) were suggested, including other neurotransmitters, neurotrophic factors, and enzymes related to the metabolism of beta amyloid protein and neuroinflammation.

### Appendix Tables

**Appendix Table 1.** Demographic Information.

|  | No exposure<br>N=12 | Recent exposure<br>N=11 | Remote exposure<br>N=12 |
| --- | --- | --- | --- |
| **Females / Males, N** | 7/5 | 6/5 | 7/5 |
| **Age, min-max years (mean)** | 28 – 64 (46.5) | 29 – 55 (41.3) | 25 – 61 (42.76) |
| **# Months in Havana, min-max (mean)** | 0 | 5 – 8 (6.5) | 1-48 (21.95) |

**Appendix Table 2.**
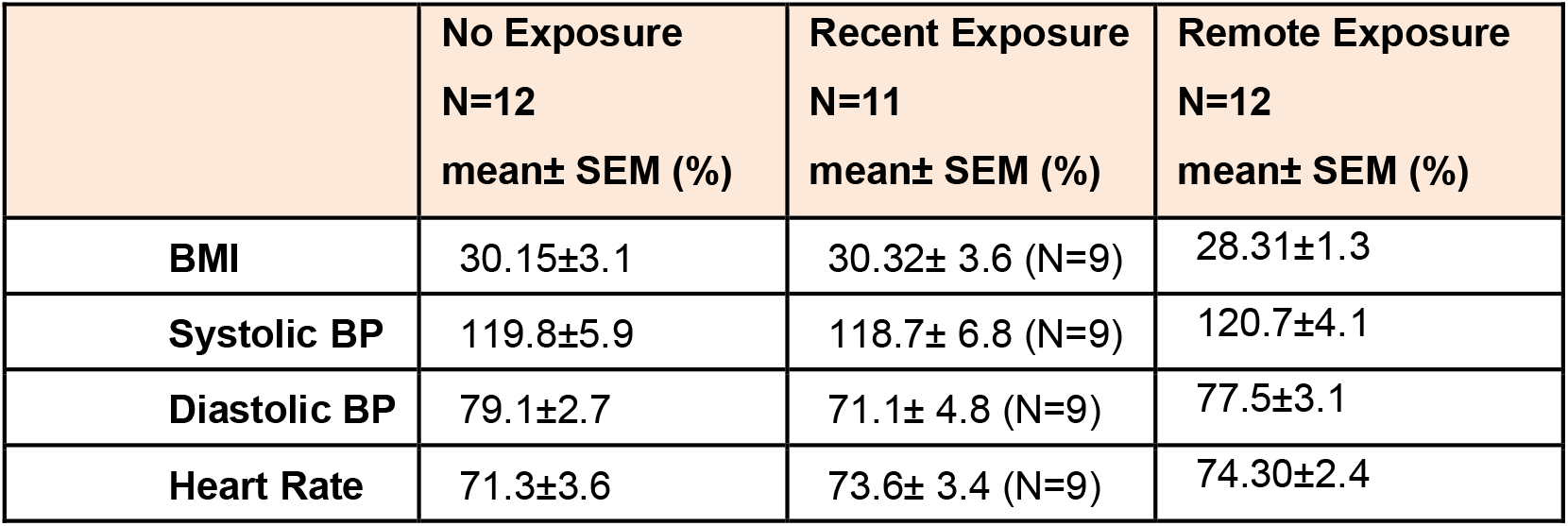
Anthropometric Measures.

|  | No Exposure<br>N=12<br>mean± SEM (%) | Recent Exposure<br>N=11<br>mean± SEM (%) | Remote Exposure<br>N=12<br>mean± SEM (%) |
| --- | --- | --- | --- |
| **BMI** | 30.15±3.1 | 30.32± 3.6 (N=9) | 28.31±1.3 |
| **Systolic BP** | 119.8±5.9 | 118.7± 6.8 (N=9) | 120.7±4.1 |
| **Diastolic BP** | 79.1±2.7 | 71.1± 4.8 (N=9) | 77.5±3.1 |
| **Heart Rate** | 71.3±3.6 | 73.6± 3.4 (N=9) | 74.30±2.4 |

**Appendix Table 3.**
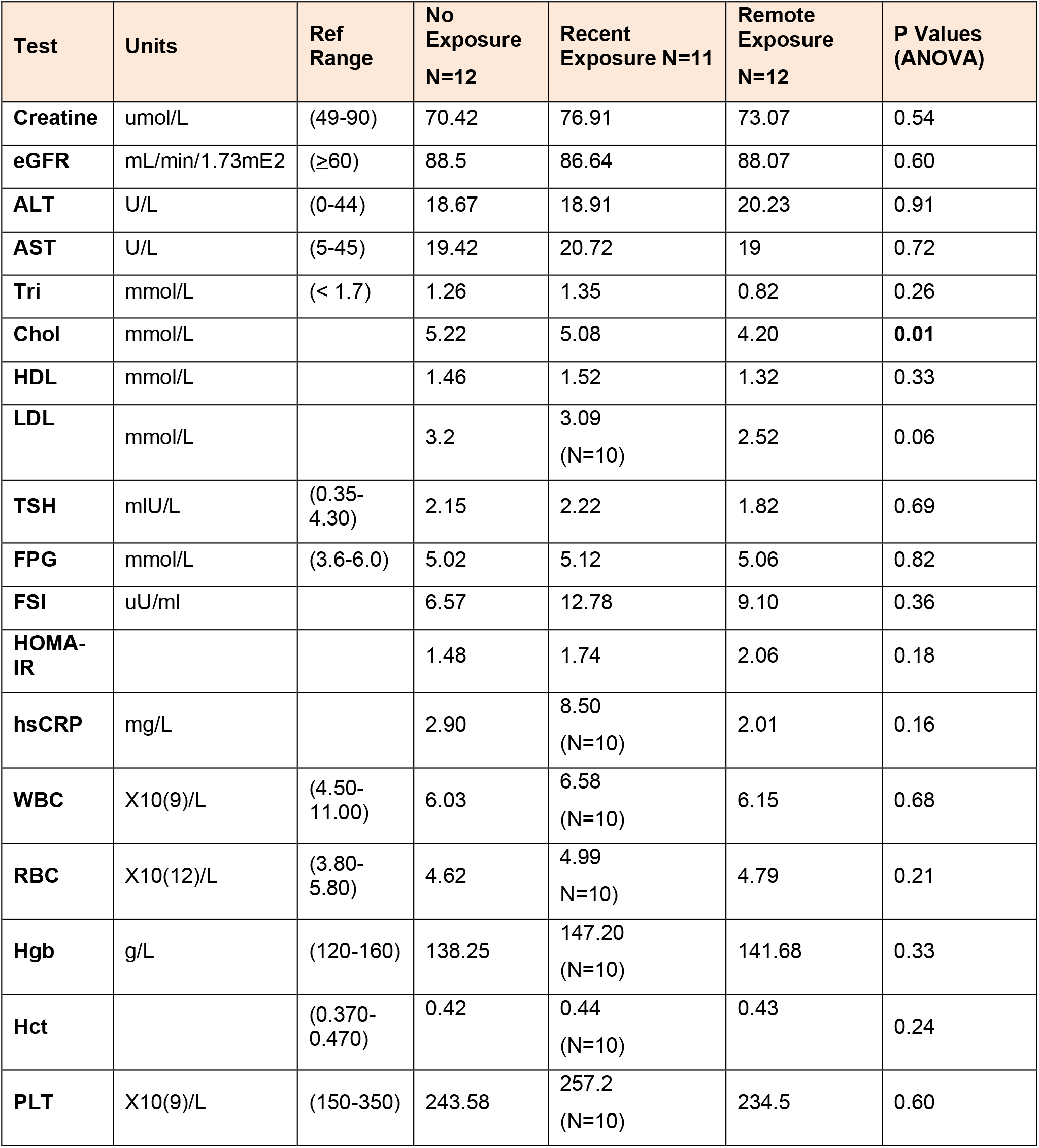
Blood Tests.

**Appendix Table 4.**
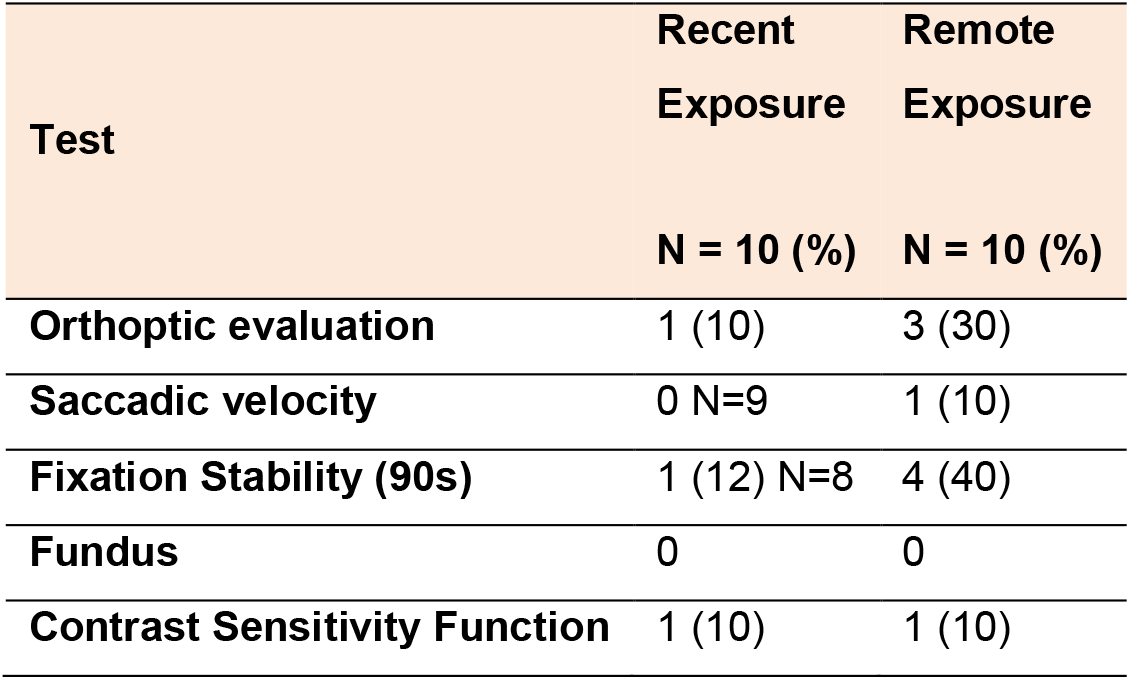
Visual Assessment Results.

**Appendix Table 5.** Auditory Assessment Results.

| Test | Recent<br>Exposure | Remote<br>Exposure | P Value<br>($\chi^2$) |
| --- | --- | --- | --- |
|  | N = 10 (%) | N = 10 (%) |  |
| ABR Interpeak Latency<br>(I-III, III-V, I-V) >2.5 SDV | 8 (80) | 6 (60) | 0.6 |
| ABR Interaural Wave V<br>> 0.3 ms | 1 (10) | 0 | 1.0 |
| Absolute latency<br>(waves I,III,V) >2.5 SDV | 6 (60) | 5 (50) | 1.0 |
| Acoustic Reflexes | 8 (80) | 8 (80) | 1.0 |

**Appendix Table 6.** Vestibular Assessment Results.

| Test | Recent Exposure<br>N = 9 (%) | Remote Exposure<br>N = 6 (%) | P Value<br>($\chi^2$) |
| --- | --- | --- | --- |
| CVEMP | 3 (33.3) | 2 (33.3) | >0.9999 |
| OVEMP | 2 (22.2) | 4 (66.6) | 0.1357 |
| Caloric & vHIT | 1 (11.1) | 0 | >0.9999 |
| Gaze Nystagmus | 0 | 2 (33.3) | 0.1429 |
| Positional | 2 (22.2) | 0 N=4 | >0.9999 |

**Appendix Table 7.** Results of Serum Toxicological Analysis.

| Analyte | Pesticide Class | No Exposure<br>N=7 (%) | Recent Exposure<br>N=11 (%) | Remote Exposure<br>N=10 (%) | P Value<br>($\chi^2$) |
| --- | --- | --- | --- | --- | --- |
| Cypermethrin | Pyrethroid | 0 (0) | 0 (0) | 0 (0) | N/A |
| Deltamethrin | Pyrethroid | 0 (0) | 0 (0) | 0 (0) | N/A |
| Permethrin | Pyrethroid | 1 (14) | 0 (0) | 0 (0) | 0.211 |
| Tilt | Triazole | 0 (0) | 0 (0) | 0 (0) | N/A |
| Temephos# | Organophosphate | 0 (14) | 1 (9.1) | 6 (54.5) | 0.012 |
| Chlorpyrifos | Organophosphate | 0 (0) | 0 (0) | 0 (0) | N/A |
| 3-PBA# | Pyrethroid metabolite | 4 (57) | 7 (70) | 6 (54.5) | 0.750 |

### Appendix Figures

**Appendix Figure 1.**
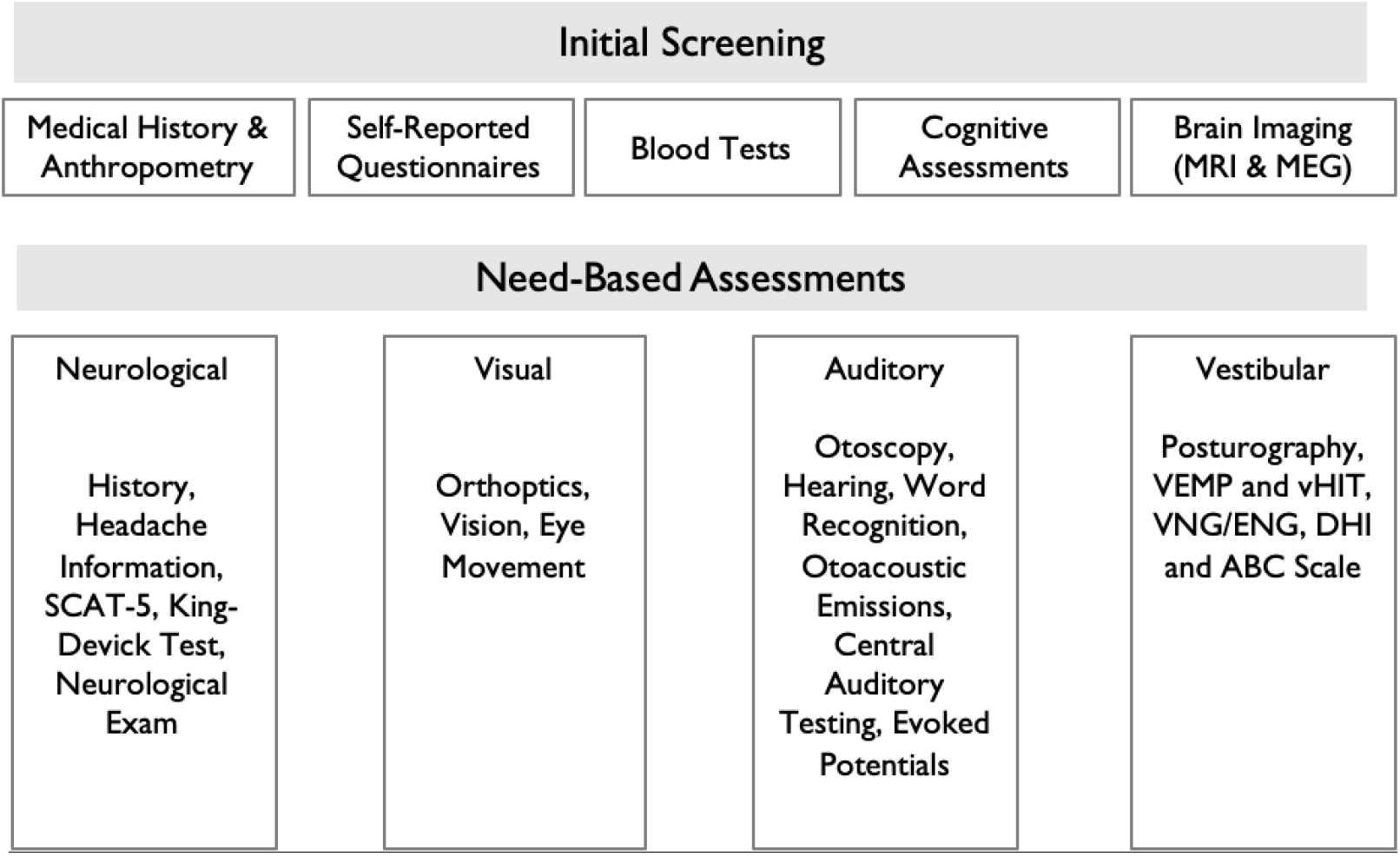
Visual Breakdown of Study Procedures. All 26 individuals underwent an initial screening (as shown) while those suspected to show abnormalities in the screening phase underwent additional need-based assessments (as shown).

**Appendix Figure 2.**
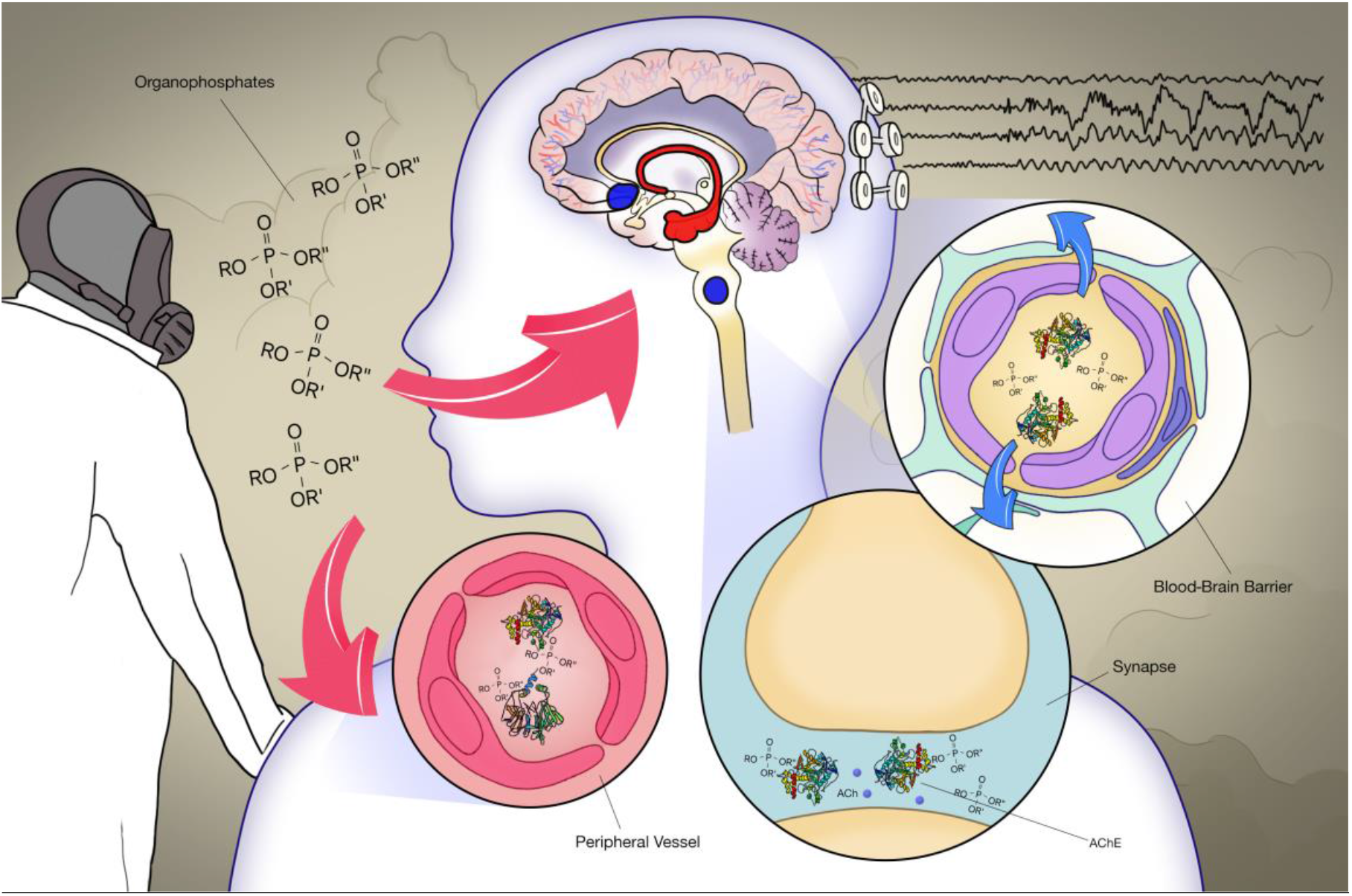
Visual Summary of Findings. Based on the clinical course, pattern of injury, and brain regions involved, we raise the hypothesis of low-dose environmental exposure to cholinesterase-inhibitors (e.g., organophosphates) as a likely cause and working medical diagnosis, with insecticides being a possible source. This narrower hypothesis (insecticides) gains contextual support from Cuba’s well-documented efforts to mitigate the spread of the Zika virus by means of mass indoor and outdoor fumigations (see Figure 5A-B in main article above),^16,17^ Canadian embassy records indicating heavy fumigation (Figure 5C), and toxicological findings in the blood (Figure 5F). We propose that cholinesterase-inhibiting organophosphates are the underlying cause for reduced cholinesterase activity in recently exposed individuals (Figure 5D-E), according to the following mechanism: organophosphates cause BBB opening and dysfunction of specific cholinergic brain centers (brainstem nuclei and basal forebrain, *marked in blue;* Figures 2-3). Consequently, axonal fibres from the basal forebrain through the fornix and to the hippocampus and parahippocampal gyri (*marked in red*) are injured (Figure 2), leading to impaired spatial memory (Appendix Table 5). Dysfunction of the basal forebrain and the related diffuse cholinergic cortical innervation underlies cortical rhythmic slowing observed in MEG (Figure 4).

**Appendix Figure 3.**
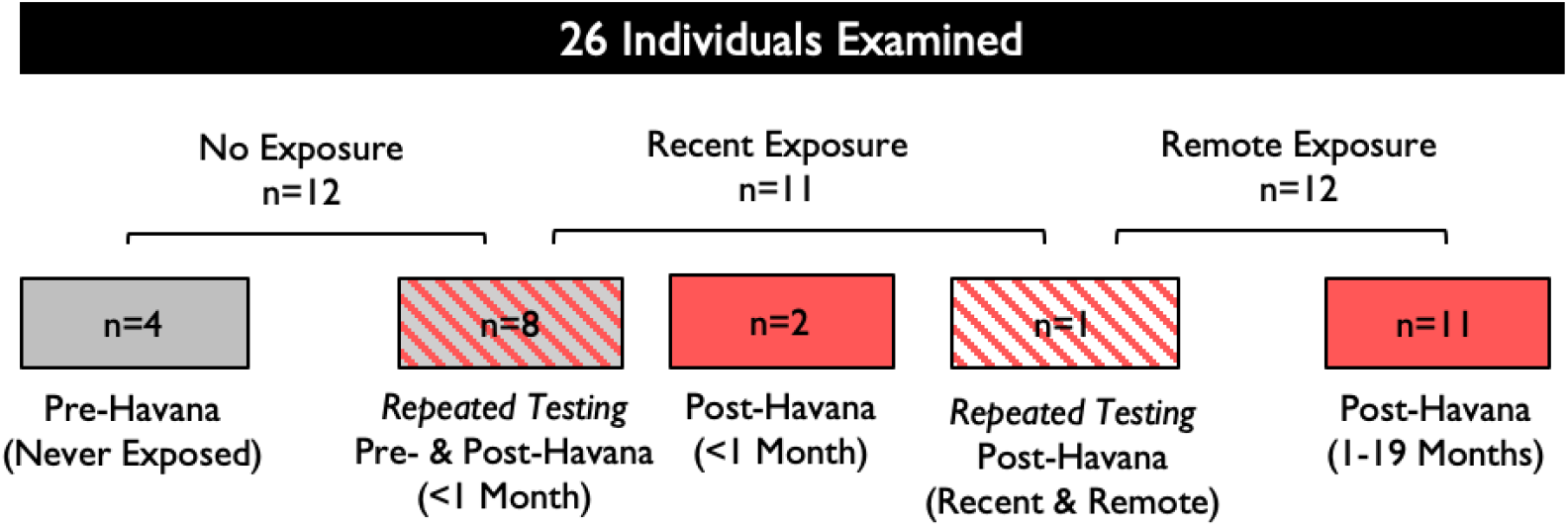
Visual Breakdown of Study Cohort. We tested 26 Canadian adult subjects for evaluation: 23 were considered “exposed” while 12 were considered “non-exposed,” with double testing (pre- and post-exposure) accounting for overlap between the two groups. Within the exposed group, we tested 11 individuals within one month of their return from Havana, a group we classified as “recently exposed,” and 12 individuals 1-19 months after returning (median 14 months), a group we classified as “remotely exposed,” with double testing accounting for overlap between these two subgroups. In particular, 8 participants were tested both prior to and immediately upon returning from Havana (forming part of both non-exposed and recent test groups), while 1 recently exposed subject was tested again as a remote subject.

## References

1 Hoffer ME, Levin BE, Snapp H, Buskirk J, Balaban C. Acute findings in an acquired neurosensory dysfunction. Laryngoscope Investig Otolaryngol 2019; 4: 124–31.

2 Swanson RL, Hampton S, Green-McKenzie J, et al. Neurological Manifestations Among US Government Personnel Reporting Directional Audible and Sensory Phenomena in Havana, Cuba. JAMA 2018; 319: 1125–33.

3 Ngwenya LB, Gardner RC, Yue JK, et al. Concordance of common data elements for assessment of subjective cognitive complaints after mild-traumatic brain injury: a TRACK-TBI Pilot Study. Brain Inj 2018; 32: 1071–8.

4 King NS, Crawford S, Wenden FJ, Moss NEG, Wade DT. The Rivermead Post Concussion Symptoms Questionnaire: a measure of symptoms commonly experienced after head injury and its reliability. J Neurol 1995; 242: 587–92.

5 Lipton RB, Stewart WF, Sawyer J, Edmeads JG. Clinical Utility of an Instrument Assessing Migraine Disability: The Migraine Disability Assessment (MIDAS) Questionnaire. Headache J Head Face Pain 2008; 41: 854–61.

6 Yang M, Rendas-Baum R, Varon SF, Kosinski M. Validation of the Headache Impact Test (HIT-6™) across episodic and chronic migraine. Cephalalgia 2011; 31: 357–67.

7 Green KL, Brown GK, Jager-Hyman S, Cha J, Steer RA, Beck AT. The Predictive Validity of the Beck Depression Inventory Suicide Item. J Clin Psychiatry 2015; 76: 1683–6.

8 Wang Y-P, Gorenstein C. Assessment of depression in medical patients: a systematic review of the utility of the Beck Depression Inventory-II. Clinics (Sao Paulo) 2013; 68: 1274–87.

9 Blevins CA, Weathers FW, Davis MT, Witte TK, Domino JL. The Posttraumatic Stress Disorder Checklist for DSM-5 (PCL-5): Development and Initial Psychometric Evaluation. J Trauma Stress 2015; 28: 489–98.

10 Buysse DJ, Reynolds CF, Monk TH, Berman SR, Kupfer DJ. The Pittsburgh Sleep Quality Index: a new instrument for psychiatric practice and research. Psychiatry Res 1989; 28: 193–213.

11 Tournier J-D, Calamante F, Connelly A. MRtrix: Diffusion tractography in crossing fiber regions. Int J Imaging Syst Technol 2012; 22: 53–66.

12 Raffelt DA, Tournier J-D, Smith RE, et al. Investigating white matter fibre density and morphology using fixel-based analysis. Neuroimage 2017; 144: 58–73.

13 Weissberg I, Veksler R, Kamintsky L, et al. Imaging blood-brain barrier dysfunction in football players. JAMA Neurol 2014; 71. DOI:10.1001/jamaneurol.2014.2682.

14 Bardouille T, Power L, Lalancette M, et al. Variability and bias between magnetoencephalography systems in non-invasive localization of the primary somatosensory cortex. Clin Neurol Neurosurg 2018; 171: 63–9.

15 Bisset JA, Rodríguez MM, Ricardo Y, et al. Temephos resistance and esterase activity in the mosquito Aedes aegypti in Havana, Cuba increased dramatically between 2006 and 2008. Med Vet Entomol 2011; 25: 233–9.

16 How Cuba is defeating the feared Zika virus. https://www.statnews.com/2016/11/08/zika-in-cuba/ (accessed April 19, 2019).

17 Reardon S. Mosquito guns and heavy fines: how Cuba kept Zika at bay for so long. Nature 2016; 536: 257–8.

18 Tumolo J. ‘Sonic Attacks’ on U.S. Diplomats in Cuba. Hear J 2019; 72: 22.

19 Dyer Montreal O. Microwave weapon caused syndrome in diplomats in Cuba, US medical team believes. DOI:10.1001/jama.2018.1742.

20 Browne RO, Moyal-Segal L Ben, Zumsteg D, et al. No Title. 2006; 20. DOI:10.1096/fj.05-5576fje.

21 Benmoyal-Segal L, Vander T, Shifman S, et al. Acetylcholinesterase/paraoxonase interactions increase the risk of insecticide-induced Parkinson’s disease. FASEB J 2005; 19: 452–4.

22 Nation DA, Sweeney MD, Montagne A, et al. Blood–brain barrier breakdown is an early biomarker of human cognitive dysfunction. Nat Med 2019; 25: 270–6.

23 Yanagisawa N, Morita H, Nakajima T. Sarin experiences in Japan: Acute toxicity and long-term effects. J Neurol Sci 2006; 249: 76–85.

24 Stone R. U.K. attack puts nerve agent in the spotlight. Science 2018; 359: 1314–5.

25 Bowers MB, Goodman E, Sim VM. SOME BEHAVIORAL CHANGES IN MAN FOLLOWING ANTICHOLINESTERASE ADMINISTRATION. J Nerv Ment Dis 1964; 138: 383–9.

26 Yamasue H, Abe O, Kasai K, et al. Human brain structural change related to acute single exposure to sarin. Ann Neurol 2007; 61: 37–46.

27 Bar-Klein G, Lublinsky S, Kamintsky L, et al. Imaging blood-brain barrier dysfunction as a biomarker for epileptogenesis. Brain 2017; 140: 1692–705.

28 del Pino J, Moyano P, Anadon MJ, et al. SN56 basal forebrain cholinergic neuronal loss after acute and long-term chlorpyrifos exposure through oxidative stress generation; P75NTR and α7-nAChRs alterations mediated partially by AChE variants disruption. Toxicology 2016; 353–354: 48–57.

29 Duffy FH, Burchfiel JL, Bartels PH, Gaon M, Sim VM. Long-term effects of an organophosphate upon the human electroencephalogram. Toxicol Appl Pharmacol 1979; 47: 161–76.

30 Zaganas I, Kapetanaki S, Mastorodemos V, et al. Linking pesticide exposure and dementia: what is the evidence? Toxicology 2013; 307: 3–11.

31 Dundar MA, Derin S, Aricigil M, Eryilmaz MA. Sudden bilateral hearing loss after organophosphate inhalation. Turkish J Emerg Med 2016; 16: 171–2.

32 Bohbot VD, Kalina M, Stepankova K, Spackova N, Petrides M, Nadel L. Spatial memory deficits in patients with lesions to the right hippocampus and to the right parahippocampal cortex. Neuropsychologia 1998; 36: 1217–38.

33 Dassanayake T, Gawarammana IB, Weerasinghe V, et al. Auditory event-related potential changes in chronic occupational exposure to organophosphate pesticides. Clin Neurophysiol 2009; 120: 1693–8.

34 França DMVR, Bender Moreira Lacerda A, Lobato D, et al. Adverse effects of pesticides on central auditory functions in tobacco growers. Int J Audiol 2017; 56: 233–41.

35 Aitken P, Zheng Y, Smith PF. The modulation of hippocampal theta rhythm by the vestibular system. J Neurophysiol 2018; 119: 548–62.

36 Garcia-Rill E, Hyde J, Kezunovic N, Urbano FJ, Petersen E. The physiology of the pedunculopontine nucleus: implications for deep brain stimulation. J Neural Transm 2015; 122: 225–35.

37 Grange-Messent V, Bouchaud C, Jamme M, Lallement G, Foquin A, Carpentier P. Seizure-related opening of the blood-brain barrier produced by the anticholinesterase compound, soman: new ultrastructural observations. Cell Mol Biol (Noisy-le-grand) 1999; 45: 1–14.

38 Craig ADB. How do you feel--now? The anterior insula and human awareness. Nat Rev Neurosci 2009; 10: 59–70.

39 McCrory P, Meeuwisse W, Dvorak J, et al. Consensus statement on concussion in sport—the 5 th international conference on concussion in sport held in Berlin, October 2016. Br J Sports Med 2017; : bjsports-2017-097699.

40 Galetta KMM, Liu M, Leong DFF, Ventura REE, Galetta SLL, Balcer LJJ. The King-Devick test of rapid number naming for concussion detection: meta-analysis and systematic review of the literature. Concussion 2016; 1: cnc.15.8.

41 Smith SM, Jenkinson M, Woolrich MW, et al. Advances in functional and structural MR image analysis and implementation as FSL. Neuroimage 2004; 23: S208–19.

42 Veksler R, Shelef I, Friedman A. Blood-Brain Barrier Imaging in Human Neuropathologies. Arch Med Res 2014; 45. DOI:10.1016/j.arcmed.2014.11.016.

43 Chassidim Y, Veksler R, Lublinsky S, Pell GSGS, Friedman A, Shelef I. Quantitative imaging assessment of blood-brain barrier permeability in humans. Fluids Barriers CNS 2013; 10: 9.

44 Taulu S, Simola J. Spatiotemporal signal space separation method for rejecting nearby interference in MEG measurements. Phys Med Biol 2006; 51: 1759–68.

45 Browne RO, Moyal-Segal L Ben, Zumsteg D, et al. Coding region paraoxonase polymorphisms dictate accentuated neuronal reactions in chronic, sub-threshold pesticide exposure. FASEB J 2006; 20: 1733–5.

46 Sweeney C, Park U, Kim JC. Comparison of sample preparation approaches and validation of an extraction method for nitrosatable pesticides and metabolites in human serum and urine analyzed by liquid chromatography - orbital ion trap mass spectrometry. J Chromatogr 2019; under revi.

47 Sánchez-Santed F, Colomina MT, Herrero Hernández E. Organophosphate pesticide exposure and neurodegeneration. Cortex 2016; 74: 417–26.

48 Pereira EFR, Aracava Y, DeTolla LJ, et al. Animal Models That Best Reproduce the Clinical Manifestations of Human Intoxication with Organophosphorus Compounds. J Pharmacol Exp Ther 2014; 350: 313–21.

49 Naughton SX, Terry A V. Neurotoxicity in acute and repeated organophosphate exposure. Toxicology 2018; 408: 101–12.

## References

18 Tumolo J. ‘Sonic Attacks’ on U.S. Diplomats in Cuba. Hear J 2019; 72: 22.

20 Browne RO, Moyal-Segal L Ben, Zumsteg D, et al. No Title. 2006; 20. DOI:10.1096/fj.05-5576fje.

24 Stone R. U.K. attack puts nerve agent in the spotlight. Science 2018; 359: 1314–5.

